# Participant perspectives to improve tenofovir adherence in the prevention of mother-to-child transmission of hepatitis B virus in Kinshasa, DRC

**DOI:** 10.1101/2023.03.30.23287808

**Authors:** Camille E. Morgan, Sahal Thahir, Patrick Ngimbi, Melchior Kashamuka Mwandagalirwa, Sarah Ntambua, Jolie Matondo, Martine Tabala, Charles Mbendi, Didine Kaba, Marcel Yotebieng, Jonathan B. Parr, Kristin Banek, Peyton Thompson

**Affiliations:** University of North Carolina at Chapel Hill, Chapel Hill, USA; Kinshasa School of Public Health, Kinshasa, Democratic Republic of Congo; University Hospital, Kinshasa, Democratic Republic of Congo; Albert Einstein College of Medicine, New York, USA

**Keywords:** pregnancy, HBV, Africa, PMTCT, tenofovir disoproxil fumarate, antenatal care

## Abstract

Prevention of mother-to-child transmission (PMTCT) programs for hepatitis B virus (HBV) are critical to reach the World Health Organization’s 2030 HBV elimination goals. Despite demonstrated feasibility utilizing HIV infrastructure, HBV PMTCT programs are not implemented in many African settings, including in the Democratic Republic of Congo (DRC). In a previous pilot of HBV PMTCT implementation in DRC’s capital, Kinshasa, we observed low TDF metabolite levels at delivery among women with high-risk HBV who were given tenofovir disoproxil fumarate (TDF) antiviral therapy. As such, we conducted qualitative interviews with women who received TDF to understand facilitators and barriers of medication adherence. We used a modified Information-Motivation-Behavioral Skills model (IMB+) as a framework for thematic content analysis. We found that trust in healthcare workers, familial support, and improved awareness of the disease and treatment options were important facilitators of TDF adherence; pill size, social stigma, and low HBV knowledge were barriers to adherence. While overall acceptance of TDF was high in this pilot, improved TDF adherence is needed in order to reach efficacious levels for preventing transmission from mothers to newborns. We suggest ongoing HBV sensitization within existing maternity and HIV care infrastructure would address gaps in knowledge and stigma identified here. Additionally, given the trust women have towards maternity center staff and volunteers, scaled HBV PMTCT interventions should include specific sensitization and education for healthcare affiliates, who currently receive no HBV prevention or information in DRC. This study is timely as TDF, particularly future long-acting formulations, could be considered as an alternate rather than adjuvant to birth-dose vaccination for HBV PMTCT in sub-Saharan Africa.

## Introduction

Prevention of mother-to-child transmission (PMTCT) is a key component of the World Health Organization (WHO)’s global HBV elimination strategy due to infants’ high exposure risk and vulnerability for developing chronic infection.(1) Current WHO guidelines for HBV PMTCT focus on timely HBV birth-dose vaccination of all infants and peripartum antiviral prophylaxis with tenofovir disoproxil fumarate (TDF) for pregnant women with HBV at high-risk of transmission (defined by high viral load and/or HBV e antigen [HBeAg] positivity).(2) While antenatal HBV screening and treatment is similar to HIV PMTCT programs, HBV is a less well-known infection(3,4) and HBV PMTCT programs are less widely implemented compared with programs for HIV.(5,6)

The Democratic Republic of Congo (DRC) currently has no implemented HBV elimination activities, apart from the three-dose infant pentavalent vaccine that includes HBV immunization. Infant HBV vaccination was first introduced in the national infant immunization program in 2007, but no catch-up vaccination programs were conducted for those born prior to roll-out nor has birth-dose vaccine been adopted. Further, healthcare workers are not routinely vaccinated against HBV, nor are they provided with ongoing bloodborne pathogen education. The existing HIV PMTCT infrastructure, which includes education about HIV, screening for HIV at the first antenatal care visit, post-test counseling by volunteer “mother mentors,” and antiretroviral therapy for women who test positive for HIV,(7) is the ideal platform for implementing an HBV PMTCT program. In the capital city of Kinshasa, where HBV prevalence among expectant mothers is estimated between 2.7-4.9%,(8,9) our group has demonstrated the feasibility and acceptability of using this existing HIV PMTCT infrastructure for HBV PMTCT implementation. Through the Arresting Vertical Transmission of HBV (AVERT-HBV) study, we implemented HBV surface antigen (HBsAg) screening alongside antenatal HIV screening, conducted HBV viral load and HBeAg testing, provided prophylaxis to women with high-risk HBV, and administered HBV birth-dose vaccination to all infants born to women with HBV.(8) We provided education and sensitization about HBV to the respective health zone staff, maternity center staff, and the volunteer mother mentors, the latter of whom serve as key points of contact for women throughout their ante- and postnatal care.

The AVERT-HBV parent study initially assessed TDF adherence via three methods: self-report, pill count and quantification of TDF’s active moiety, tenofovir diphosphate (TFVdp), in dried blood spots (DBS) collected at delivery. All nine women given TDF reported daily adherence and presented empty pill bottles, but only one of the nine women given TDF had TFVdp levels reflective of full adherence. These findings prompted follow-up qualitative interviews with mothers with high-risk HBV in the study who received TDF prophylaxis to learn about their experience with the medication. We sought to collect participants insights into facilitators and barriers to TDF adherence, to inform roll-out of TDF prophylaxis within future HBV elimination policy in DRC.

To contextualize this study, we conducted a literature search of TDF adherence for HBV, in which we found no studies of adherence to antiviral prophylaxis for HBV in pregnancy. Few studies have analyzed the social and psychological facilitators of adherence to HBV antiviral therapy within chronic treatment programs; and none specific to pregnant women or to contexts in Africa.(10,11) In the few studies of adherence within chronic treatment programs, urban residence, non-cirrhotic status, and family member reminders predicted high adherence,(10) and adherence tends to be higher with tenofovir-based regimens than with other antivirals.(11) None of these studies applied a behavioral model to identify barriers and facilitators of adherence to antivirals systematically.

Our study in Kinshasa, DRC addresses this absence of research on TDF adherence for HBV, which is critical to inform scale-up of HBV elimination activities in a region with high HBV mortality and incidence and few prevention measures widely implemented.(12) Through this analysis, we provide detailed recommendations for strengthening antiviral adherence for expanded HBV PMTCT programs in Kinshasa, DRC, now the largest city in Africa and one of the fastest growing among them.(13)

## Materials and Methods

### Setting

This study was conducted within the Arresting Vertical Transmission of HBV (AVERT-HBV) study in two high-volume maternity centers in Kinshasa, DRC, (8) a city of 16 million people and with no widely implemented HBV PMTCT programs. This parent study of 90 mother-infant dyads utilized existing infrastructure for HIV PMTCT by incorporating: 1) HBV sensitization to maternity center staff and participants, 2) rapid HBsAg screening alongside existing rapid HIV screening, 3) HBV viral load and HBV envelope antigen (HBeAg) testing, 4) peripartum tenofovir disoproxil fumarate (TDF) therapy of women at high risk of HBV transmission (viral load ≥200,000 IU/mL or HBeAg-positivty), and 5) monovalent HBV birth-dose vaccination for all infants.(8) Pregnant women who were HBsAg-positive, older than 18 years of age, ≤24 weeks’ gestation, healthy without requiring hospitalization during pregnancy, and intending to continue maternity and post-partum care at these respective maternity centers, were enrolled in the AVERT-HBV study. Women with HBV of high-risk of transmission were provided TDF prophylaxis, following WHO HBV PMTCT guidelines.(14) A total of nine women received the once-daily TDF regimen prior to delivery (starting between 28 and 32 weeks of gestation) through 12 weeks post-partum; adherence was initially evaluated through pill counts and measurement of TDF’s active moiety, tenofovir diphosphate (TFVdp), in DBS samples. All nine were newly diagnosed with HBV during the original study, and none tested positive for HIV. Using DBS collected at the delivery visit, we observed only one woman to have a level of TFVdp reflective at least 4-7 doses taken per week. We initiated qualitative interviews to explore the participants’ experience with the medication to inform HBV PMTCT implementation in this setting.

### Data collection and analysis

As the subgroup who received TDF prophylaxis was small (n=9), we sought to interview all women. Study authors (PN, JM, SN) initiated phone interviews with all women with high-risk HBV who received TDF prophylaxis and conducted interviews with all who were reachable by phone. Interviews occurred approximately six to nine months after study completion, and lasted 45-60 minutes. Interviews were conducted in French or Lingala based on the woman’s preference. After additional verbal consent was provided, the women were asked broadly about their experiences with study enrollment, perceived benefits and side effects of tenofovir, factors affecting their medication adherence, and social impacts of their new HBV diagnosis. The complete interview guide can be found in the supplementary materials (S-1). The research team recorded, transcribed, and translated interviews from Lingala to French and then to English.

### Theoretical framework and analysis

The information-motivation-behavior (IMB) model has been shown to be an effective theoretical foundation for assessing HIV-related adherence behaviors in both high- and low-resource settings,(8,15– 17) and is ideal for adapting to a HBV PMTCT program that utilized existing HIV infrastructure.(8) Fisher *et al*. first proposed the IMB model, outlining three constructs: (1) *Information* includes pertinent medication knowledge including administration, side effects, and broader understanding of adherence-related outcomes; (2) *Motivation* includes personal and social influences on medical adherence; (3) *Behavioral skills* includes the individual’s objective and perceived capacity to complete adherence-related tasks.(18) For the analysis, an updated version of the IMB model (IMB+) which describes how personal or contextual factors may influence the different IMB constructs was used.(15) Using thematic content analysis, three team members (CEM, ST, KB) coded the data by identifying IMB+ themes and other patterns across interviews. Results were discussed and aggregated into key themes by study team consensus.

### Ethical considerations

This study was approved by the Institutional Review Board at the University of North Carolina at Chapel Hill (IRB# 17-2090) and the Kinshasa School of Public Health (CE/ESP/001/2018). Written informed consent had been received from women in the parent study, and verbal consent was received for telephone interviews and recordings at the beginning of the interview. Participant IDs have been changed in this text of this manuscript from the ones used in the course of the study and are only known to research team members.

## Results

### Participant characteristics

Six of the nine women who received TDF prophylaxis were reachable by telephone and were interviewed. Demographic and clinical characteristics of the six women were similar to those not able to be reached (**Supplementary Tables 1-2**). The ages of participating women ranged from 19 to 35 years with a median age of 25 years. Gravidity ranged from one to eight with a median of two. Five of six women were married or in a marriage-like relationship, half had received post-secondary education, and one third reported employment at enrollment in the parent study.

#### The IMB+ model for tenofovir adherence

Responses from the six participants who shared their experiences taking TDF were mapped to show how information, motivation, and behavior skills influenced TDF adherence. All participants discussed personal and contextual factors beyond the traditional IMB model that affected their adherence; these are categorized as modulating factors (**Figure 1**).

**Figure 1.**
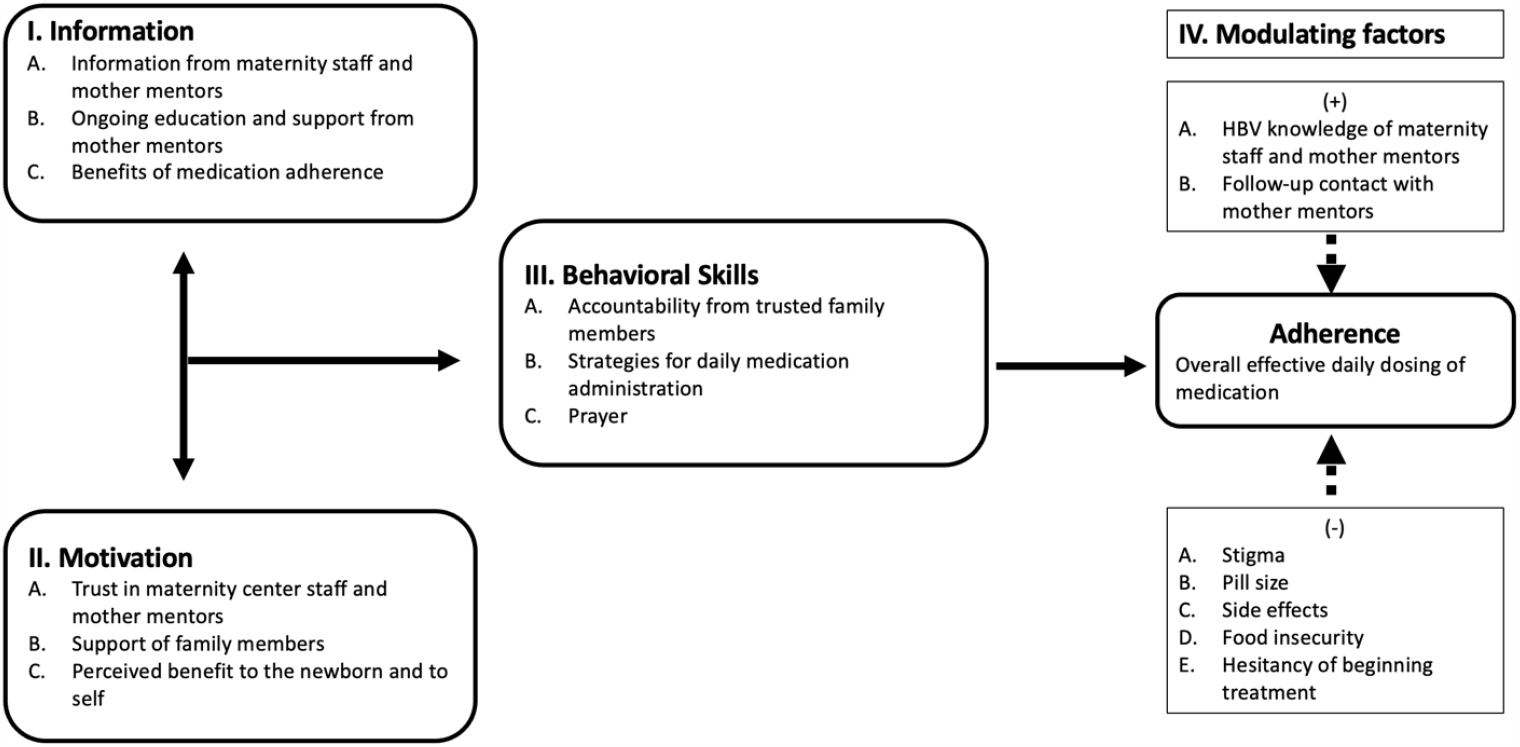
A modified Information-Motivation-Behavioral Skills (IMB+) model for analysis of tenofovir adherence among pregnant women in the DRC. The positive or negative influences of these factors are indicated by plus and minus signs, respectively.

#### Information

The information component of the IMB model for TDF adherence was represented in the participants’ understanding of the HBV disease process, the treatment regimen, and the benefit of treatment (**Figure 1**). Many of the participants highlighted that the first time they had heard of HBV was at their diagnosis upon study enrollment. Participants frequently cited receiving information about HBV and TDF from maternity center representatives at the initial and follow-up visits (**Table 1, A)**, as well as ongoing support from mother mentors (**Table 1, B**).

**Table 1.**
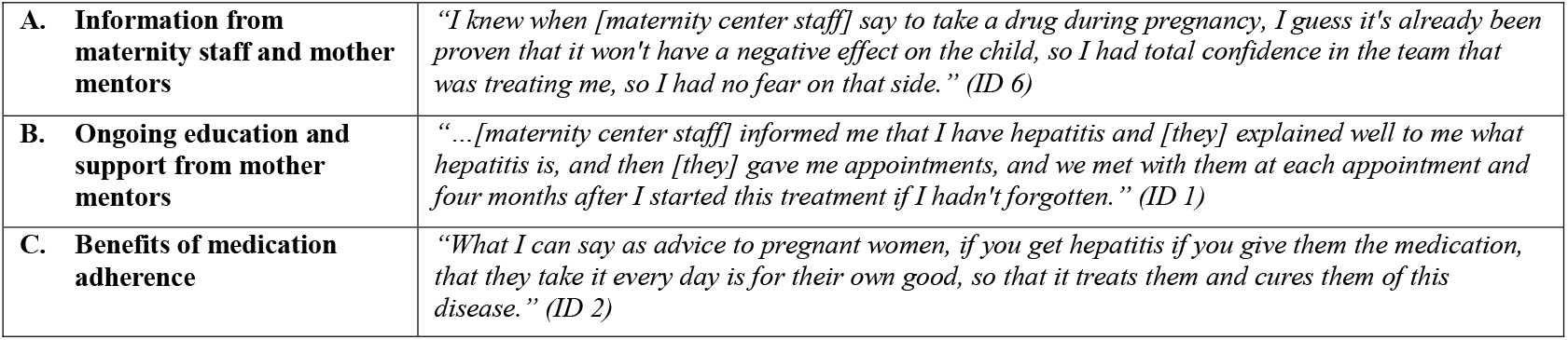
Representative quotes for the Information component of the IMB+ analysis from interviews with six mothers with high-risk HBV.

|  |  |
| --- | --- |
| **A. Information from maternity staff and mother mentors** | *"I knew when [maternity center staff] say to take a drug during pregnancy, I guess it's already been proven that it won't have a negative effect on the child, so I had total confidence in the team that was treating me, so I had no fear on that side." (ID 6)* |
| **B. Ongoing education and support from mother mentors** | *"...[maternity center staff] informed me that I have hepatitis and [they] explained well to me what hepatitis is, and then [they] gave me appointments, and we met with them at each appointment and four months after I started this treatment if I hadn't forgotten." (ID 1)* |
| **C. Benefits of medication adherence** | *"What I can say as advice to pregnant women, if you get hepatitis if you give them the medication, that they take it every day is for their own good, so that it treats them and cures them of this disease." (ID 2)* |

### Motivation

Participants discussed the social influences and their personal incentives motivating their TDF adherence. A key theme was overall trust in the healthcare team and the information they received from maternity staff compared with circulating information (**Table 2, A.1**). Another participant also demonstrated the same sentiment of trust by discussing how her trust in the medical team helped spur her adherence to TDF during the study (**Table 2, A.2**).

**Table 2.**
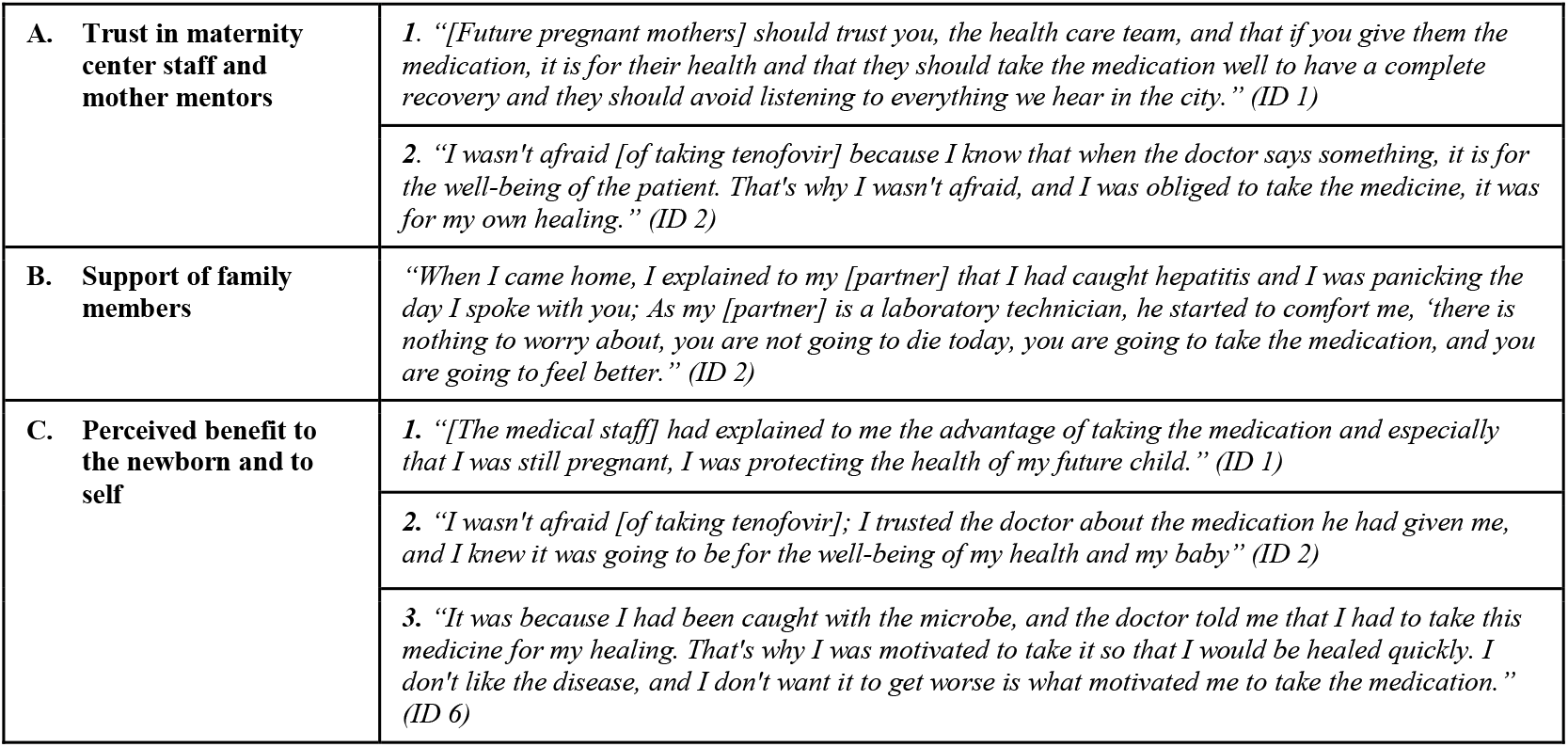
Representative quotes for the Motivation component of the IMB+ analysis from interviews with six mothers with high-risk HBV.

|  |  |
| --- | --- |
| **A. Trust in maternity center staff and mother mentors** | *1. "[Future pregnant mothers] should trust you, the health care team, and that if you give them the medication, it is for their health and that they should take the medication well to have a complete recovery and they should avoid listening to everything we hear in the city." (ID 1)* |
|  | *2. "I wasn't afraid [of taking tenofovir] because I know that when the doctor says something, it is for the well-being of the patient. That's why I wasn't afraid, and I was obliged to take the medicine, it was for my own healing." (ID 2)* |
| **B. Support of family members** | *"When I came home, I explained to my [partner] that I had caught hepatitis and I was panicking the day I spoke with you; As my [partner] is a laboratory technician, he started to comfort me, 'there is nothing to worry about, you are not going to die today, you are going to take the medication, and you are going to feel better.'" (ID 2)* |
| **C. Perceived benefit to the newborn and to self** | *1. "[The medical staff] had explained to me the advantage of taking the medication and especially that I was still pregnant, I was protecting the health of my future child." (ID 1)* |
|  | *2. "I wasn't afraid [of taking tenofovir]; I trusted the doctor about the medication he had given me, and I knew it was going to be for the well-being of my health and my baby" (ID 2)* |
|  | *3. "It was because I had been caught with the microbe, and the doctor told me that I had to take this medicine for my healing. That's why I was motivated to take it so that I would be healed quickly. I don't like the disease, and I don't want it to get worse is what motivated me to take the medication." (ID 6)* |

An additional social motivation for adherence was the emotional support of family members, specifically when participants shared their new diagnosis. Many participants shared how family members encouraged them at the time of diagnosis. One participant shared how her initial interactions with her partner eased her anxiety about her new diagnosis and encouraged her TDF adherence (**Table 2, B**). Internal motivation for medication adherence was primarily defined by participants’ perceived positive outcomes for their unborn children and themselves: (**Table 2, C**).

### Behavioral Skills

Women described strategies to assist them in taking the daily TDF dose, including prayer, social accountability, and visual cues (**Figure 1**). One participant shared her status and treatment plan with her sibling who lives with her, in order to have accountability in taking the medication (**Table 3, A**). This same participant described the routine she developed after speaking with the maternity representatives to help ease her anxiety when taking the medication (**Table 3, C)**. Another participant described placing the medication bottle by her child’s stories, a place where she would be reminded to take it (**Table 3, B**).

**Table 3.**
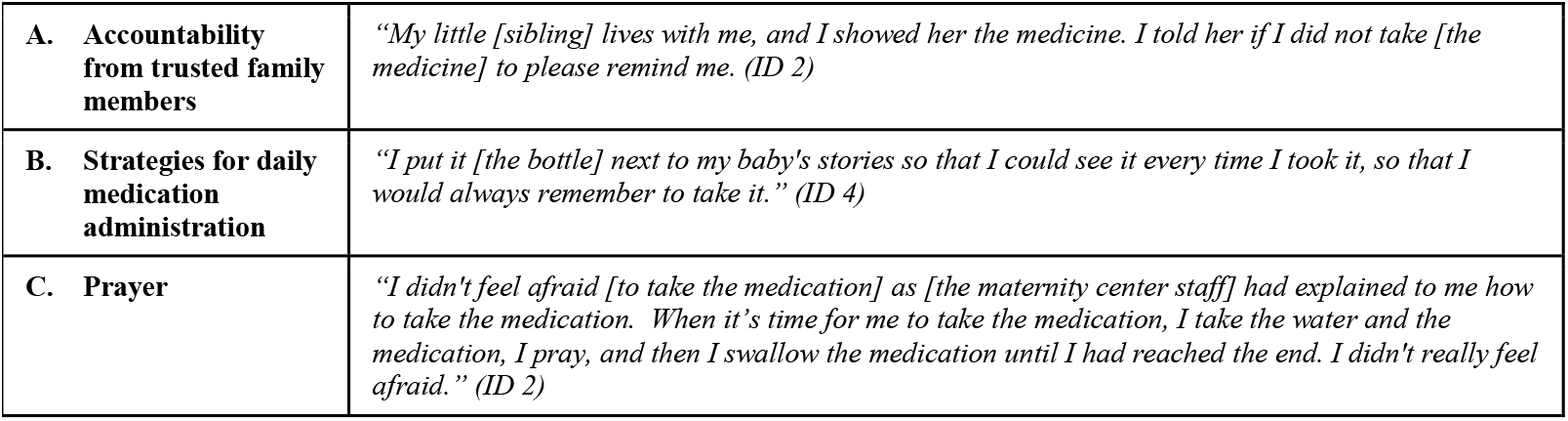
Representative quotes for the Behavioral Skills component of the IMB+ analysis from interviews with six mothers with high-risk HBV.

### Modulating Factors

#### Positive influences

Women reported the information they continued to receive from maternity representatives as contributing to their adherence. Volunteer mother mentors serve not only as resources with information about HBV, but also as reminders for following the treatment regimen. The longitudinal nature and trust of these relationships contributed positively to TDF adherence (**Table 4, A-B)**.

**Table 4.** Representative quotes for Modulating Factors component of the IMB+ analysis from interviews with six mothers with high-risk HBV.

|  |  |  |
| --- | --- | --- |
| (+) | **A. Knowledge of maternity center staff and mother mentors** | *"...the first day I was given the medication, I felt embarrassed with a fear that people would find out that I had the disease, but after some explanations from [maternity center staff] I had finally gotten rid of that embarrassment." (ID 2)* |
|  | **B. Follow-up contact with mother mentors** | *"...at times I also forgot to take the medication for three days, but when I saw that she kept giving me the bottle of medication all month long, I said to myself, 'Good, as I am taking their medication, I think it's a reality and that maybe with this medication this disease will be able to end, that's when I became aware of it, and I began to respect the medication.'" (ID 4)* |
| (-) | **C. Stigma** | *1. "I just didn't want people to see my medication to avoid too many questions about it, knowing my environment, people always talk badly. That's why I didn't want other family members to see me with this medication....Yes, in fact, there was a time when I didn't come to the center for follow-up, [the study nurse] had sent someone from the center to pick me up, and when the man arrived at my house, he met my [relative], and I stayed with her, and this [relative] was able to turn the message into saying that I have HIV all over the [place], and it had weakened me a lot, and I was very worried when I took the medication, but then I didn't care about that, I remained objective when I took my medication until I was cured." (ID 6)* |
|  |  | *2. "...the first day I was given the medication, I felt embarrassed with a fear that people would find out that I had the disease, but after some explanations from [maternity center staff] I had finally gotten rid of that embarrassment." (ID 2)* |
|  | **D. Pill size** | *"At the beginning of the treatment, I forgot to take it. I was taking it with difficulty because the tablets were large." (ID 4)* |
|  | **E. Side effects** | *"I felt a little uncomfortable, and when I took this medication I felt tired, sleepy and especially as I was still big [pregnant] and I would need a little rest, I would have to sleep for a few minutes, and when I woke up I could feel much better until I had given birth I kept on taking it, so it was more fatigue, but for a while, I came back strong, all that was just the effects of the medication." (ID 2)* |
|  | **F. Food insecurity** | *"Yes, I have missed days to take the medication. It's hunger, sometimes I missed something to eat, and as you know the medicine with hunger doesn't work. When you take the medication on an empty stomach, you will feel very bad." (ID 3)* |
|  | **G. Hesitancy of beginning treatment** | *"Yes, a little bit of forgetting, but that didn't happen several times. I took my medication every day. At first, I tended to forget, but as time went by, when I became familiar with this medication, I took it like paracetamol. I don't remember how many times I missed taking the medication, but it wasn't much, I think it was only 3-4 days. [The reason I didn't take the medication every day] is forgetting, and also I didn't have the courage to take the medication at the beginning of the treatment." (ID 6)* |

#### Negative influences

A main modifying factor that emerged from the data was the barrier of social stigma related to HBV. A participant discussed the stigma and embarrassment she felt while taking the medication, leading to a missed follow-up appointment at the maternity center and further rumors. Despite this social influence, she reported maintaining TDF adherence (**Table 4, C1**).

Medication characteristics and side effects were also mentioned as factors influencing treatment adherence. One participant mentioned that she initially had difficulty with the pill size, and another described fatigue in her experience with taking the medication (**Table 4, D-E**). One woman reported a lack of having food as a reason for missing doses, as she perceived taking medication on an empty stomach would make her feel bad (**Table 4, F**). Initial hesitancy and forgetfulness about the medication was also cited as a reason for not taking all doses, which improved with time (**Table 4, G)**.

## Discussion

This qualitative study explored facilitators and barriers to TDF adherence for preventing mother-to-child transmission of hepatitis B in the Democratic Republic of Congo. Highlighting the experiences and voices of participants is essential for context-specific understanding of adherence to inform future scale-up of HBV elimination efforts.(19)

A key facilitator of adherence for individuals in this study was trust in maternity center staff and mother mentors, along with the education these individuals provided about HBV and TDF prophylaxis. Having supportive patient-provider relationships and high trust in health workers improved patient access to services, cooperation, and adherence to medications in other studies.(20–22) Trust in health workers has also been shown to improve the acceptance and effectiveness of community-based maternal-child health initiatives,(23) especially adherence to antiretrovirals in PMTCT programs.(24) It is worth noting distinct aspects of the antenatal care context in which this program was conducted: women in this study built rapport with mother mentors over the course of routine obstetric care, as these volunteers not only provide initial counseling, but also conduct follow-up; if a woman failed to return for subsequent antenatal care visits for HIV care, or in our case, for HBV care, it was the mother mentor who followed up by phone or in person on behalf of our research team. The established trust that women have in mother mentors will be beneficial for HBV PMTCT program scale-up, and their role should be prioritized in future program design.

Findings also highlighted the resiliency women had following their diagnoses of HBV. While there is limited literature on HBV treatment adherence, HIV literature has demonstrated that factors including optimism, self-efficacy, spirituality/religiousness, and social support, all of which were exhibited by participants in this study, are key to resiliency in ART adherence.(25) Participants shared the vital role social support played in ensuring medication adherence. These interviews reflected the uncertainty and fear women experienced following their diagnoses, and the support they sought from their family members or the healthcare staff at the maternity centers. Such social support has been observed to improve adherence in one of few studies of TDF adherence in a chronic HBV treatment program,(10) as well as in adherence to HIV treatment.(26–29)

We observed pill size, social stigma, and limited baseline knowledge about HBV and treatment were barriers to optimal TDF adherence. Medication characteristics, such as pill size and quantity, are noted by the US Food and Drug Administration as established barriers for medication adherence;(30,31) we did observe these barriers eased over time once the participants became accustomed to the treatment, which suggests these barriers can be addressed with ongoing education and continuity of care. Stigma is also a known impeding factor to seeking care and adhering to treatment for HBV within chronic treatment programs.(32) In our study, women cited fear that people around them would misconceive HBV as HIV, a more widely known and stigmatized infection.(33) As with pill size, the fear of stigma appeared to decrease for the women in this study the longer they took the treatment. This finding is similar to the effects of stigma on tuberculosis (TB) treatment adherence. Treatment for TB is similar to HBV prophylaxis for PMTCT in its finite course, and reassuringly, social stigma does not appear to decrease adherence to TB treatment consistently.(34,35)

Overall, study participants had little knowledge of prevention, transmission, or symptoms of HBV, consistent with previous findings among pregnant women in this setting and similar ones,(36–38) and in broader sub-Saharan African settings.(39–41) Beliefs, misguided or not, can affect adherence,(42) but are generally found to be less effective than improved knowledge in the disease process and therapy.(43,44) A critical misguided belief observed in this study was that the treatment would cure the women of HBV. While some women reported this as a motivating factor for medication adherence, this belief reflects a broader misunderstanding of HBV disease. Similar misunderstandings have been documented as reasons for poor adherence to ART in studies of HIV treatment adherence.(45) Our results also highlighted other misconceptions that worsened adherence, including the belief by one woman that food intake was required with medication. While food intake is not required for the medication,(46) this misconception led to poorer adherence. Food insecurity has been described as a factor decreasing ART adherence in people living with HIV, which is likely to apply in HBV treatment adherence as well.(47)

The recall, selection, and social desirability biases are worth noting as study limitations. First, we conducted these interviews six to nine months after completing the parent study, which was 12 to 14 months after completion of their TDF treatment courses, inclusive of a delay due to the COVID-19 pandemic. Such a lag in time may have led to recall bias, omitting or exaggerating factors influencing TDF adherence. Additionally, while we sought to interview all women with high-risk HBV who received TDF, the women seeking antenatal care at the two study maternity centers in the parent study might not represent women seeking care at other maternity hospitals in Kinshasa. Further, participants might have provided responses that they perceived to be what the study team wanted to hear; we believe social desirability bias here is limited by the fact that the study interviewers were maternity nurses with whom participants had strong, longitudinal relationships that fostered candor from participants. Lastly, while six interviewees could be considered a small sample size, the consistency of themes across participants suggests that saturation was reached; the demographic and clinical similarity between the six women and the three not interviewed suggests these interviews may represent the women with high-risk HBV from the parent study. Given the lack of data relating to TDF adherence for HBV PMTCT, these findings offer important perspectives for future research and clinical programs despite these biases.

## Conclusions

As scarce research has been conducted on HBV treatment adherence for PMTCT, these findings from participant interviews provide specific recommendations for HBV PMTCT policy and programs in low-resourced settings. HBV PMTCT programs may have more successful adherence and thus prevention if education campaigns target pregnant mothers, mother mentors, and maternity center staff. Sensitization about HBV can address low knowledge as well as stigma. Given the trustworthy resource that mother mentors and maternity center staff are to pregnant women, these individuals are important to include as recipients and educators in ongoing awareness efforts. HIV sensitization is already implemented at antenatal clinics in this setting, offering the ideal platform for integrating HBV education to accompany HBV PMTCT programs. Future implementation research should focus on design of educational programming, to determine the most efficacious approach for TDF adherence and thus successful prevention of HBV transmission. Lastly, this study is timely as antiviral prophylaxis is being evaluated as an alternative option to HBV birth-dose vaccination in sub-Saharan Africa.

## Data Availability

The datasets generated and/or analyzed during the current study are not publicly available because they contain protected health information but are available from the corresponding author on reasonable request.

## Declarations

### Ethics approval

The study was conducted according to the guidelines of the Declaration of Helsinki, and approved by the Institutional Review Board at the University of North Carolina at Chapel Hill (IRB# 17-2090) and the Kinshasa School of Public Health (CE/ESP/001/2018).

### Informed consent

Written informed consent was obtained from all subjects for the parent study. Verbal informed consent was obtained prior to each telephone interview. There are no identifying images or information in this publication.

### Consent for publication

The authors of this submission accept the conditions of submission and the BMV Copyright and License Agreement.

## Authors’ contributions

PT, PN, KM, and JP designed the study. PN, JM, and SN collected the data. ST, CM, and KB analyzed the data. ST, CM, KB and PT wrote the first draft. All authors approved the final draft.

## Acknowledgements

We thank all of the women who participated in this study, the staff at the Binza and Kingasani maternity clinics, and provincial and national health authorities. We would like to thank Dr. Noro Ravelomanana, and Dr. Bienvenue Kawende for their contributions to the parent AVERT study.

We are grateful for the support we have received from the administrative staff at the University of North Carolina and at Kinshasa School of Public Health. We grieve the loss of Dr. Steven Meshnick, who had a major role in this study and whose vision and mentorship were critical to its success.

## List of abbreviations

ART: anti-retroviral therapy
AVERT: Arresting Vertical Transmission of Hepatitis B Virus in the DRC (parent study)
DRC: Democratic Republic of Congo
HAART: highly active anti-retroviral therapy
HBV: Hepatitis B virus
HBeAg: Hepatitis B virus e antigen
HBsAg: Hepatitis B virus surface antigen
IMB: Information-Motivation-Behavioral Skills model
PMTCT: Prevention of mother-to-child transmission
TDF: tenofovir disoproxil fumarate
TFVdp: tenofovir diphosphate

